# Effectiveness of *attexis*, a Digital Intervention Based on Cognitive Behavioral Therapy for Adults with ADHD: A Randomized Controlled Trial

**DOI:** 10.1101/2025.08.27.25334574

**Authors:** Roberto D’Amelio, Linda T. Betz, Sarah M. Jow, Wolfgang Retz, Alexandra Philipsen, Jan Philipp Klein, Eva Faßbinder, Gitta A. Jacob, Petra Retz-Junginger

## Abstract

**Background:** Access to evidence-based psychosocial interventions for adults with Attention-Deficit/Hyperactivity Disorder (ADHD) remains limited, despite strong patient demand for non-pharmacological options such as cognitive behavioral therapy (CBT). Digital interventions may offer a scalable, low-threshold solution to meet this need and complement existing care.

**Methods:** This pragmatic randomized controlled trial evaluated the effectiveness of *attexis*, a fully self-guided digital intervention based on CBT and mindfulness principles, as an adjunct to treatment as usual (TAU). A total of 337 adults with confirmed ADHD were randomized to either *attexis* + TAU or TAU alone. The primary outcome was ADHD symptom severity (Adult ADHD Self-Report Scale total score) at 3 months post-randomization (T1). Secondary outcomes included functional impairment, depressive symptoms, self-esteem, and health-related quality of life. Follow-up was conducted at 6 months (T2).

**Results:** Intent-to-treat analyses showed significantly lower ADHD symptom severity in the intervention group at T1 (baseline-adjusted mean difference = –5.0 points; *d* = 0.85, *p* < .001). Significant improvements were also observed across all secondary outcomes, and effects remained stable at T2. Responder analyses confirmed the clinical relevance of the findings. Subgroup analyses demonstrated consistent effects across sex, medication use, psychotherapy status, and treatment changes. No adverse events related to *attexis* were reported.

**Conclusions:** *attexis* was effective in reducing ADHD symptoms and improving a broad range of functional and psychosocial outcomes. As a safe, low-threshold, fully self-guided intervention, it may serve as a valuable adjunct to routine care and help address existing gaps in access to psychosocial treatment for adults with ADHD.

## Introduction

Attention-Deficit/Hyperactivity Disorder (ADHD) affects between 2.5% and 6.8% of adults globally, with an estimated average prevalence of 3.3% in high-income countries (Fayyad et al., 2017; Simon, Czobor, Bálint, Mészáros, & Bitter, 2009; Song et al., 2021). In Germany, however, estimates based on retrospective claims data from statutory health insurance are substantially lower, at just 0.7%, suggesting marked and longstanding underrecognition of adult ADHD (Libutzki et al., 2019; Philipsen & Döpfner, 2020; Schlander, Schwarz, Trott, Viapiano, & Bonauer, 2007). This is particularly concerning given the high burden of the disorder: Adults with ADHD often face substantial difficulties affecting multiple domains of adult life, including academic and occupational functioning, management of everyday responsibilities such as finances, and interpersonal relationships (de Zwaan et al., 2012; Fuermaier et al., 2021; Koerts et al., 2021). These difficulties are often accompanied by a reduced quality of life extending well into older adulthood (Thorell, Holst, & Sjöwall, 2019) as well as psychological comorbidities such as depression and anxiety, further exacerbating personal and societal burden (Faraone et al., 2024; Fayyad et al., 2017; Libutzki et al., 2019).

Current European clinical guidelines emphasize a multimodal approach to treating adult ADHD, combining pharmacological and psychosocial interventions to address both core symptoms as well as associated functional and psychosocial impairments (AWMF, 2017; Kooij, Bijlenga, et al., 2019; NICE, 2018). The German evidence- and consensus-based guideline (AWMF, 2017) recommends psychosocial interventions particularly in cases of insufficient response to pharmacotherapy, contraindications to medication, poor tolerability, or patient decision against pharmacological treatment. Additional indications include adults later in life needing support with diagnosis acceptance and treatment initiation, as well as cases of mild symptomatology where psychosocial treatment is considered sufficient to address low to moderate functional impairments (AWMF, 2017).

Among available psychosocial interventions, psychoeducation and cognitive-behavioral therapy (CBT) have demonstrated effectiveness in reducing ADHD symptom severity and associated disorder burden (Hoxhaj et al., 2018; Knouse, Teller, & Brooks, 2017; Liu, Hua, Lu, & Goh, 2023; Nimmo-Smith et al., 2020; Philipsen et al., 2015; Young, Moghaddam, & Tickle, 2020). Accordingly, clinical guidelines endorse both approaches, with psychoeducation recommended as the basis of standard care (AWMF, 2017; Kooij, Bijlenga, et al., 2019). However, access to qualified care in Germany remains limited: clinicians with expertise in adult ADHD are scarce (Schlander et al., 2007; Schneider, Schöttle, Hottenrott, Gallinat, & Moritz, 2023), and waiting times for psychotherapy exceed several months (Kruse et al., 2024). As a result, approximately 40% of adults with ADHD receive no psychotherapy within the first year after diagnosis, and among those who do, 30% receive only probatory sessions (Libutzki et al., 2019).

Digital interventions have demonstrated robust effectiveness across various psychiatric conditions (Andersson, 2016; Karyotaki et al., 2021; Pauley, Cuijpers, Papola, Miguel, & Karyotaki, 2023) and are therefore considered a promising strategy to help bridge the treatment gap in adult ADHD. To date, five small-scale randomized controlled trials (RCTs) have evaluated the effectiveness of digital interventions in reducing core ADHD symptom severity, reporting effect sizes between *d* = 0.42 and 1.21 (Kenter, Gjestad, Lundervold, & Nordgreen, 2023; Moëll, Kollberg, Nasri, Lindefors, & Kaldo, 2015; Nasri et al., 2023; Pettersson, Söderström, Edlund-Söderström, & Nilsson, 2017; Selaskowski et al., 2022). Despite growing evidence for their effectiveness, digital interventions have not yet been integrated into the routine care of adult ADHD in Germany, and no program is currently approved for reimbursement by statutory health insurance under the *Digital Health Application* (DiGA) scheme.

Against this background, we developed *attexis*, a fully self-guided digital intervention grounded in CBT and mindfulness principles. Thus, *attexis* requires no professional oversight and is accessible online at the user’s convenience. The program provides personalized psychoeducation and therapeutic exercises tailored to individual needs and preferences. Features such as adaptive content delivery, automated reminders, and progress tracking are integrated to support user engagement and motivation.

The goal of the present pragmatic RCT was to evaluate the effectiveness of *attexis* when used as an adjunct to existing treatment pathways. To this end, we compared *attexis* added to Treatment as Usual (TAU) to a TAU-only control group. TAU reflects the heterogeneous and unstandardized care typically received by adults with ADHD in routine practice in Germany, including pharmacological and non-pharmacological interventions, as well as no formal treatment. The primary endpoint was the severity of ADHD symptoms after 3 months. In addition, we assessed secondary endpoints reflecting functional and psychosocial outcomes relevant to patients, including functional impairment, depressive symptoms, self-esteem, and quality of life. We hypothesized that participants receiving *attexis* in addition to TAU would show greater improvements across these outcomes compared to those receiving TAU alone. Additional follow-up data were collected at 6 months to evaluate the durability of treatment effects.

## Methods

### Procedures

The study “Researching the Effectiveness of *attexis*, a Digital Health Application for Adults with Attention Deficit Hyperactivity Disorder – a Randomized Controlled Trial (READ-ADHD)” was approved by the Ethics Committee of the Medical Chamber Hamburg (reference number 2023-101052-BO-ff), and all procedures complied with the Declaration of Helsinki and General Data Protection Regulation.

Prior to recruitment, the study was registered in an international clinical trials registry (ClinicalTrials.gov ID: NCT06221930). German-speaking participants were recruited from March 2024 to June 2024 via an online advertising campaign using targeted Google Ads to reach adults with ADHD symptoms. Interested individuals were directed to a dedicated study website containing detailed study information and an option to register their interest.

After providing electronic informed consent, participants completed an initial online screening survey. Those who screened positively were invited to a structured diagnostic interview conducted by telephone. Trained psychological staff administered the German version of the DIVA-5 (Kooij, Francken, Bron, & Wynchank, 2019) to confirm an ADHD diagnosis and the Mini-DIPS (Margraf, Cwik, Pflug, & Schneider, 2017) to assess relevant current psychiatric comorbidities. Ambiguous cases were reviewed in regular supervision meetings with GAJ. Final eligibility was determined by the study physician SMJ following a comprehensive review of inclusion and exclusion criteria.

Eligible participants were randomized in a 1:1 ratio to either the intervention group (*attexis* + TAU) or the control group (TAU only), using an automated and concealed allocation system using computer-generated random numbers (simulating a digital coin toss) to perform simple randomization. Allocation was triggered automatically by the study platform upon completion of eligibility verification. Due to the nature of the pragmatic design, participants were not blinded to group allocation. As all outcomes were collected via self-report, no outcome assessors were involved, and blinding was therefore not applicable. Participants in the intervention group received immediate access to *attexis*, while control group participants were offered access upon study completion. The study was designed as a pragmatic trial, aiming to approximate real-world conditions without restricting concurrent treatment options: TAU permitted participants in both groups to continue, modify, or discontinue any concurrent treatments, including psychotherapy and pharmacotherapy. Planned treatment changes in the upcoming 3 months were an exclusion criterion at baseline, in order to better attribute observed effects to *attexis*. Unplanned treatment changes that occurred during the course of the study were allowed and documented, reflecting the pragmatic nature of the trial.

Participants completed outcome assessments online at 3 months (T1, primary time point for evaluating effectiveness) and 6 months (T2, follow-up to assess the durability of effects over time). All study procedures were conducted fully remotely; questionnaires online and diagnostic interviews by telephone. Participants received a €10 gift voucher for every completed follow-up assessment.

The required sample size was determined a priori based on the primary outcome; see Supplementary Method 1 for details.

### Inclusion and exclusion criteria

Participants of all sexes and genders were eligible for inclusion if they were between 18 and 65 years of age, and had a diagnosis of ADHD confirmed via the structured diagnostic interview DIVA-5. To meet the symptom severity threshold, participants were required to score ≥ 17 on either the inattention or the hyperactivity/impulsivity subscale of the Adult ADHD Self-Report Scale v1.1 (ASRS), in line with thresholds used in prior research on digital interventions for ADHD (Kenter et al., 2023; Moëll et al., 2015). Additional inclusion criteria were stable treatment status (including psychotherapy, medication, or no treatment) for at least 30 days prior to enrollment, sufficient knowledge of the German language, and provision of informed consent.

Participants were excluded if they had a lifetime diagnosis of a severe psychiatric disorder, including severe affective disorder, psychotic disorder, autism spectrum disorder, borderline or antisocial personality disorder, substance use disorder, or acute suicidality. Individuals were also excluded if they planned to change their current treatment regimen (e.g., psychotherapy or medication) within the 3 months following enrollment.

### Intervention

*attexis* is a self-guided online intervention based on cognitive-behavioral and mindfulness principles, developed to support adults with ADHD. *attexis* combines psychoeducation, therapeutic exercises, and personalized techniques tailored to the user’s reported needs and preferences (see Table 1). A central feature is its use of “simulated dialogues”, in which users interact with brief text passages by selecting response options that guide the flow of information and tailor the experience (see Supplementary Figure for example screenshots). In this context, *attexis* specifically addresses the interpersonal needs of patients with ADHD. For instance, depending on the individual’s attentional state, it may offer to repeat certain topics or suggest humorous or unexpected exercises to help sustain attention. This format supports personalization and sustained engagement. The program also includes simple homework tasks, flexible pacing, and automated reminders to encourage continued use. In addition to its core dialogues, *attexis* provides supplementary materials such as guided audio exercises, PDF worksheets, and summary sheets. Users may also opt to receive motivational messages via SMS or email, and self-monitoring tools are included to support behavioral tracking and progress monitoring over time.

**Table 1.** Content of *attexis*.

| Topic | Content Focus |
| --- | --- |
| **Psychoeducation** | Comprehensive information on ADHD diagnosis, symptomatology, and associated challenges; introduction to the neurodiversity perspective to foster self-understanding and reduce internalized stigma. |
| **Mindfulness** | Use of mindfulness to more quickly identify and interrupt problems related to attention and impulsivity; in this context, mindfulness is also practiced repeatedly as part of the program. |
| **Problem Identification** | Guided exploration of individual difficulties and life areas affected by ADHD to personalize the therapeutic approach and increase relevance. |
| **Stigma & Gender-Specific Aspects** | Discussion of public and internalized stigma, as well as gender-specific manifestations of ADHD and their implications for self-image and care-seeking behavior. |
| **Attention** | Analysis of individual “attention disruptors” and “attention sustainers”; techniques to enhance focus and compensate for inattention in daily life. |
| **Impulsivity & Emotion Regulation** | Development of strategies to interrupt impulsive actions; promotion of physical activity for emotional regulation and behavioral stabilization. |
| **Problem Solving** | Training in structured approaches to problem-solving using techniques such as SMART goal setting and the Eisenhower matrix. |
| **Self-Esteem & Self-Image** | Processing of negative childhood experiences related to ADHD; resource-oriented reframing of ADHD traits; use of imagery-based techniques to strengthen a positive self-concept. |
| **Resource Orientation** | Use of positive psychology principles and exercises to identify personal strengths and apply them effectively in everyday life situations. |

### Measures

Baseline data, as well as follow-up data at 3 months (T1) and 6 months (T2) post-randomization, were collected via a secure and encrypted online survey platform (*LimeSurvey*). All measures were based on self-report. Participants were invited to complete each assessment via email, and non-responders received up to three reminders. For follow-up assessments, additional contact methods for reminders, including SMS and telephone calls, were employed to maximize response rates.

#### Primary Endpoint

The primary endpoint was the ASRS total score, a widely used 18-item patient-reported outcome measure (PROM) assessing core ADHD symptoms (Kessler et al., 2005), evaluated at T1 (3 months post-randomization). The validated German version of the ASRS has demonstrated good psychometric properties in adults with ADHD (Mörstedt, Corbisiero, & Stieglitz, 2016).

#### Secondary Endpoints

Secondary outcomes included: (1) functional impairment, assessed using the Work and Social Adjustment Scale (WSAS) (Heissel et al., 2021; Lundqvist et al., 2024; Mundt, Marks, Shear, & Greist, 2002); (2) depressive symptoms, measured by the Patient Health Questionnaire-9 (PHQ-9) (Hennig, Koglin, Schmidt, Petermann, & Brähler, 2017; Kliem et al., 2024; Martin, Rief, Klaiberg, & Braehler, 2006); (3) self-esteem, assessed with the Rosenberg Self-Esteem Scale (RSES) (Masuch, Bea, Alm, Deibler, & Sobanski, 2019; Rosenberg, 1965; Roth, Decker, Herzberg, & Brähler, 2008; von Collani & Herzberg, 2003); and (4) quality of life, measured by the Assessment of Quality of Life - 8 Dimensions (AQoL-8D) (Richardson, Iezzi, Khan, & Maxwell, 2014; Richardson, Khan, Iezzi, & Maxwell, 2012).

#### Comorbidities and user satisfaction

Relevant sections from the Mini-DIPS were used to assess current psychiatric comorbidities in the sample (Margraf et al., 2017). To assess satisfaction with *attexis,* participants were asked to rate how likely they were to recommend *attexis* to a friend or colleague on a numeric rating scale from 0 (not likely at all) to 10 (extremely likely). Moreover, subjective improvement of ADHD symptoms as well as subjective improvement in the impact of ADHD on daily activities in the last 3 months (from T0 to T1 and from T1 to T2) was evaluated with the Patient Global Impression of Change scale (Guy, 1976).

#### Adverse Events

Given that *attexis* is a CE-marked Class I medical device, it is inherently associated with a very low risk profile. Nonetheless, adverse events were monitored throughout the study and operationalized as any unplanned or emergency outpatient or inpatient treatment reported for the preceding 3 months. Participants were asked to report such events at each follow-up assessment (T1 and T2). In addition, any events potentially related to the use of the intervention were documented as adverse device effects. All reported events were reviewed for potential causal association with *attexis*.

### Statistical Analyses

All analyses followed a pre-specified analysis plan and were conducted using *R*, version 4.4.1 (R Core Team, 2024). The primary analyses applied the intent-to-treat (ITT) principle under a missing at random assumption, using ANCOVA adjusted for baseline values. Results are reported as baseline-adjusted mean differences with 95% confidence intervals (CI) and standardized effect sizes (Cohen’s *d* based on estimated marginal means). Missing data were handled via bootstrapped multiple imputation (von Hippel & Bartlett, 2021). As a sensitivity analysis, we applied jump-to-reference (J2R) imputation, a conservative method assuming intervention dropouts follow the control trajectory (Bartlett, 2023). Per-protocol (PP) analyses, defined as including only intervention participants who had registered to use *attexis*, alongside all participants in the control group, applied the same statistical procedures as the ITT analyses. All analyses were repeated at T2 to assess the durability of effects. Clinical relevance was evaluated using responder analyses. For the primary endpoint, responders were defined as participants who showed a ≥ 30% reduction in the ASRS total score from baseline to T1 (Buitelaar, Montgomery, & van Zwieten-Boot, 2003). For secondary endpoints, responder criteria were based on predefined minimal clinically important differences (MCID), or on the reliable change index (RCI) where no MCID was available. Group differences in responder rates were analyzed using χ² tests and odds ratios (OR). As a safety analysis, we assessed symptom worsening, defined as any increase in the ASRS total score from baseline to T1. Subgroup analyses assessed potential treatment moderators (sex, psychotherapy, psychotropic medication, treatment changes). Statistical tests were considered significant at *p* < .05 (two-sided). A gatekeeping testing strategy was applied to control for multiplicity. Full details of the statistical analysis are provided in Supplementary Method 2.

## Results

### Recruitment and retention overview

A total of 2,058 individuals were screened for eligibility, of whom 337 met all inclusion criteria and were randomized to the intervention group (n = 164) or the control group (n = 173) (see Figure 1). The slight imbalance in group sizes is consistent with expected variation under simple randomization. Attrition rates at 3 months were 9.1% in the intervention group and 3.5% in the control group; at 6 months, attrition increased slightly to 13.4% and 8.1%, respectively. Following the predefined criteria for inclusion in the PP analyses, 163 out of 164 participants (99.4%) in the intervention group had activated the voucher to use *attexis*. The resulting PP dataset comprised 336 participants: 163 in the intervention group and all 173 in the control group.

**Figure 1.**
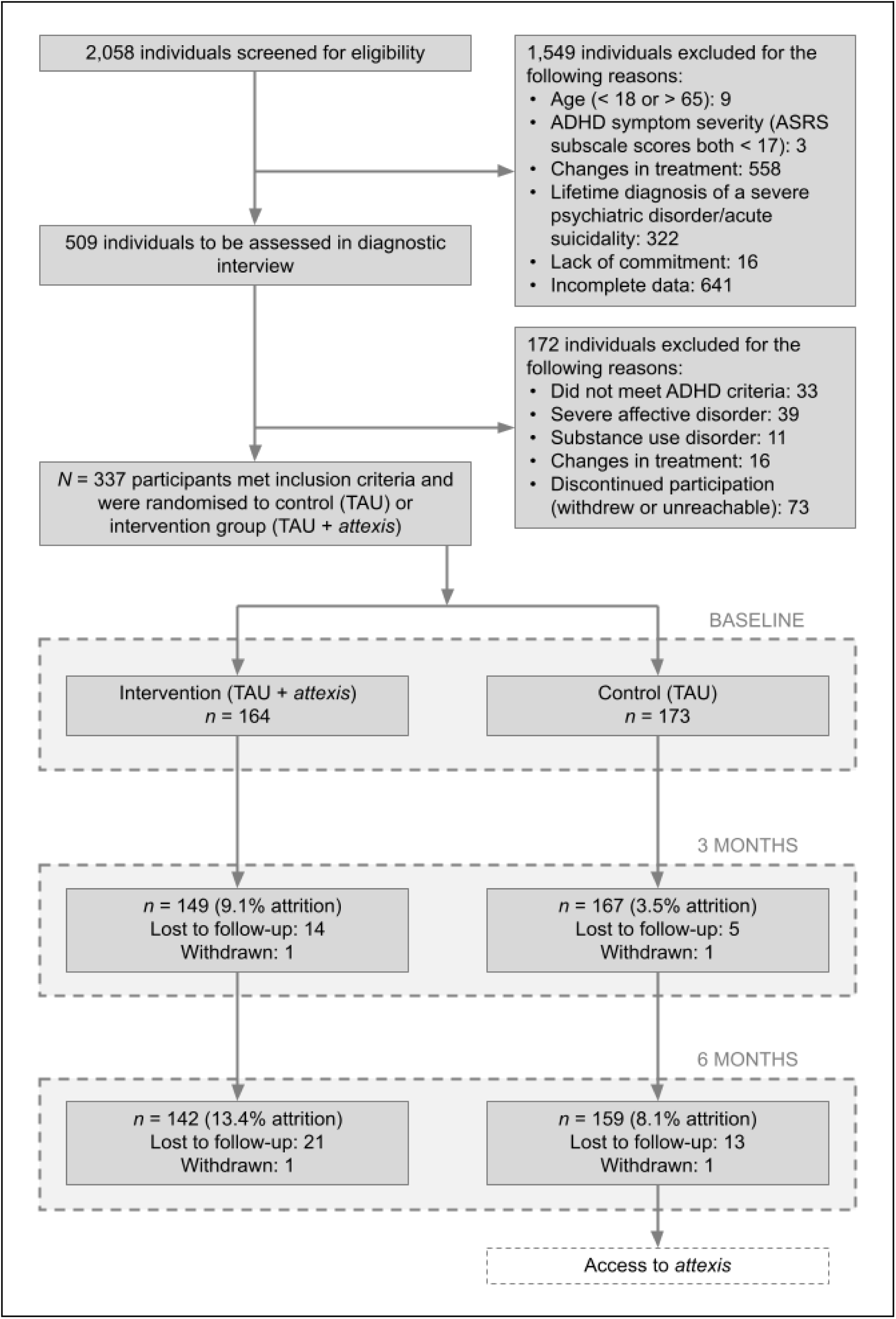
Flow of participants through the study. *Abbreviations*: ASRS = Adult ADHD Self-Report Scale; TAU = Treatment as usual.

### Baseline characteristics

Table 2 presents baseline characteristics of the study sample. The mean age was approximately 38 years, with 71% reporting female sex. Most participants were in a relationship and had a higher level of education; over half held a university degree. Employment was common, with 42% working full-time and 33% part-time. The majority identified as White (93%), with smaller proportions reporting Middle Eastern (4.5%), Latin American (2.1%), or Black (1.2%) ethnicity. Current comorbid psychiatric conditions were frequent: 27% met diagnostic criteria for major depressive disorder and 12% for social anxiety disorder. About 24% were currently in psychotherapy, and 36% were taking psychotropic medication, most commonly psychostimulants (28%).

**Table 2.**
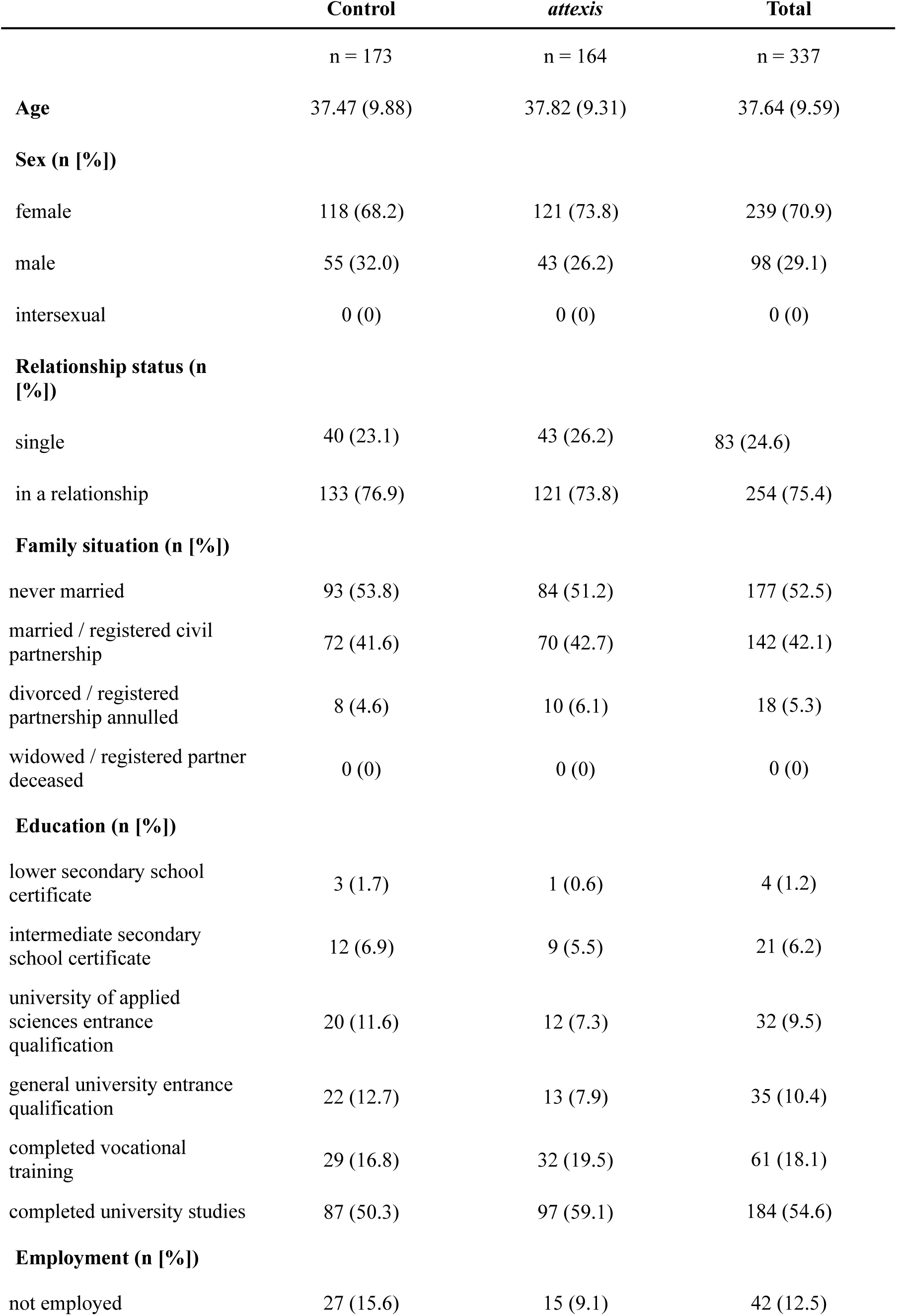
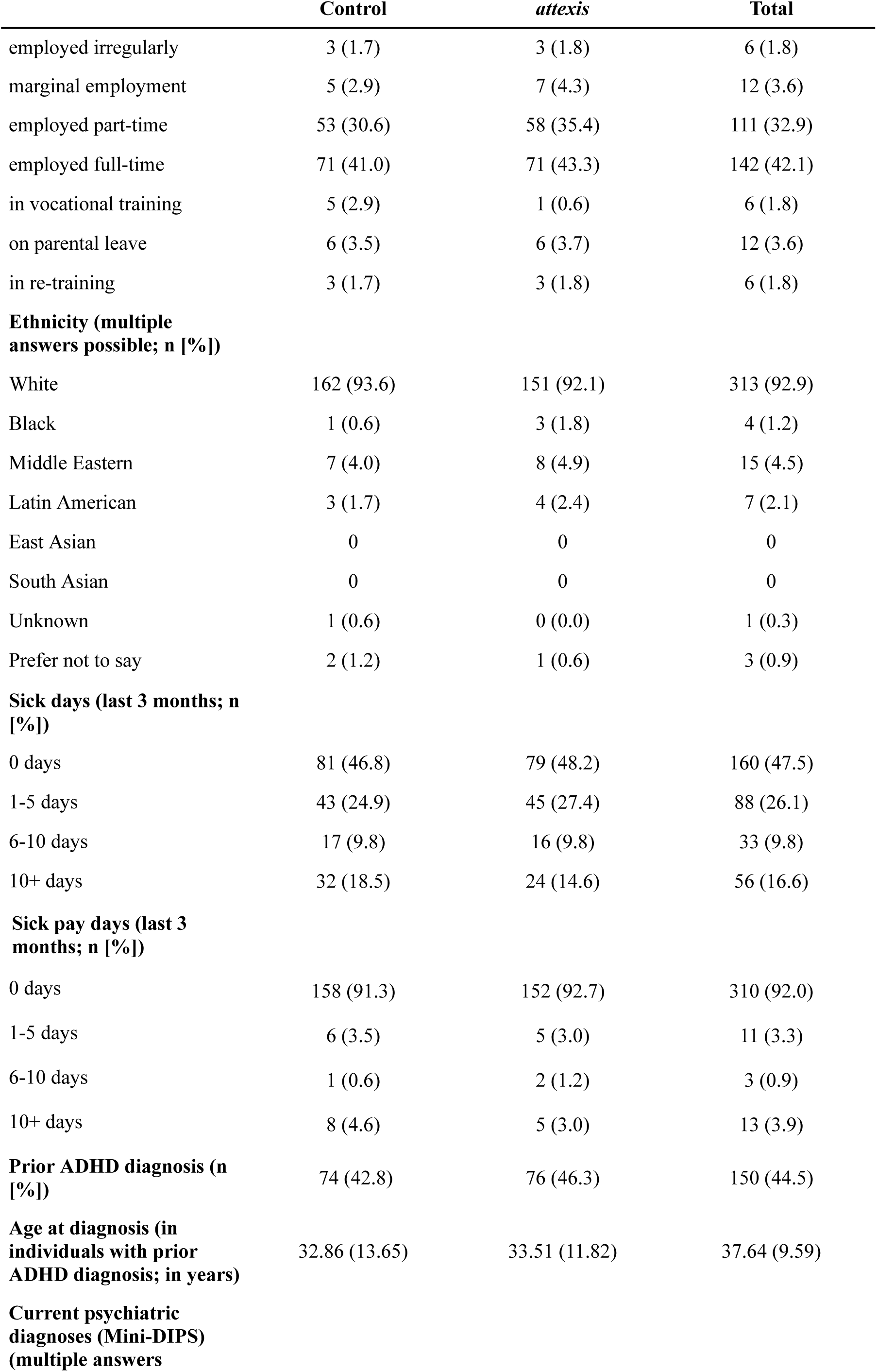
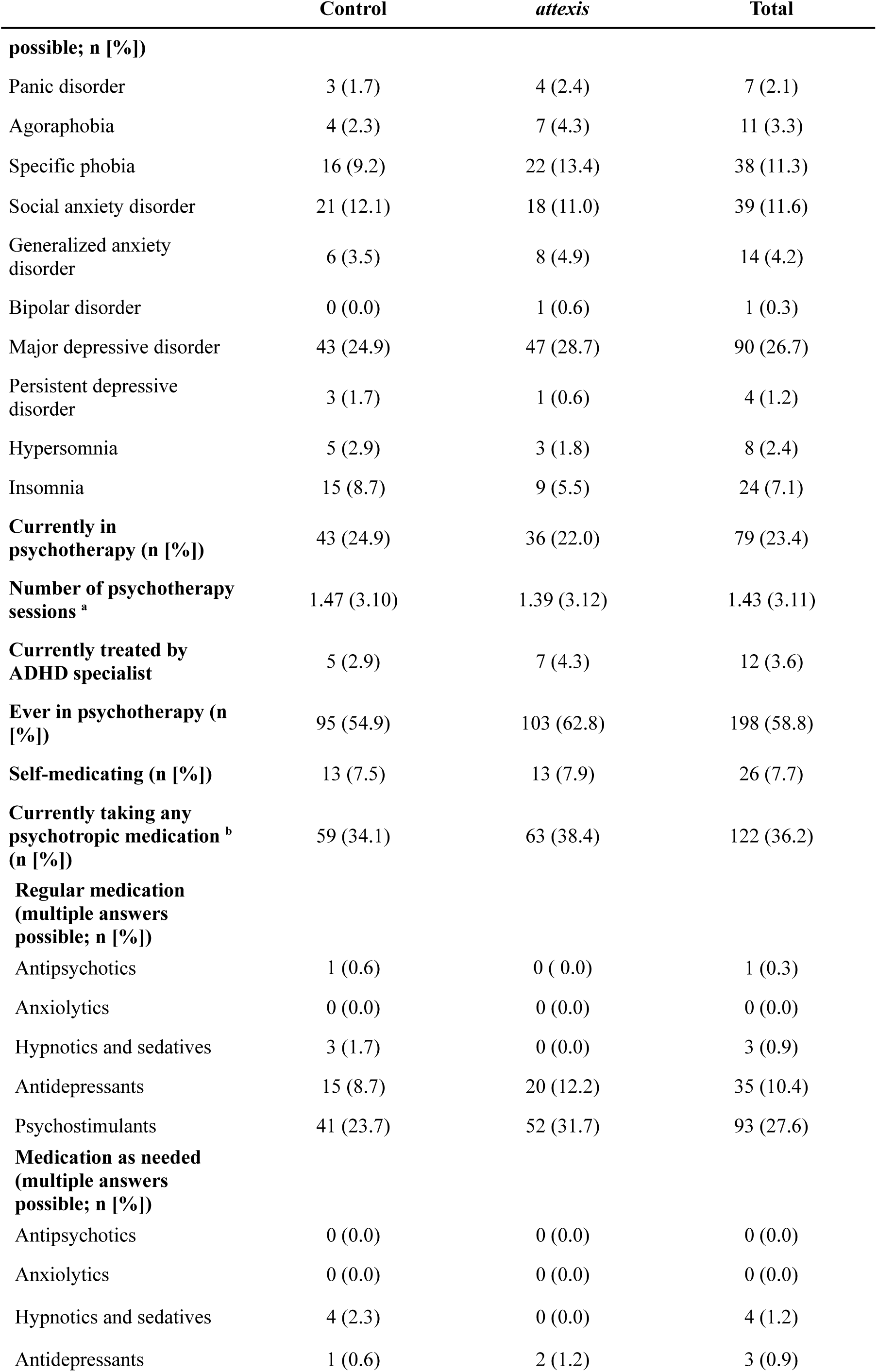
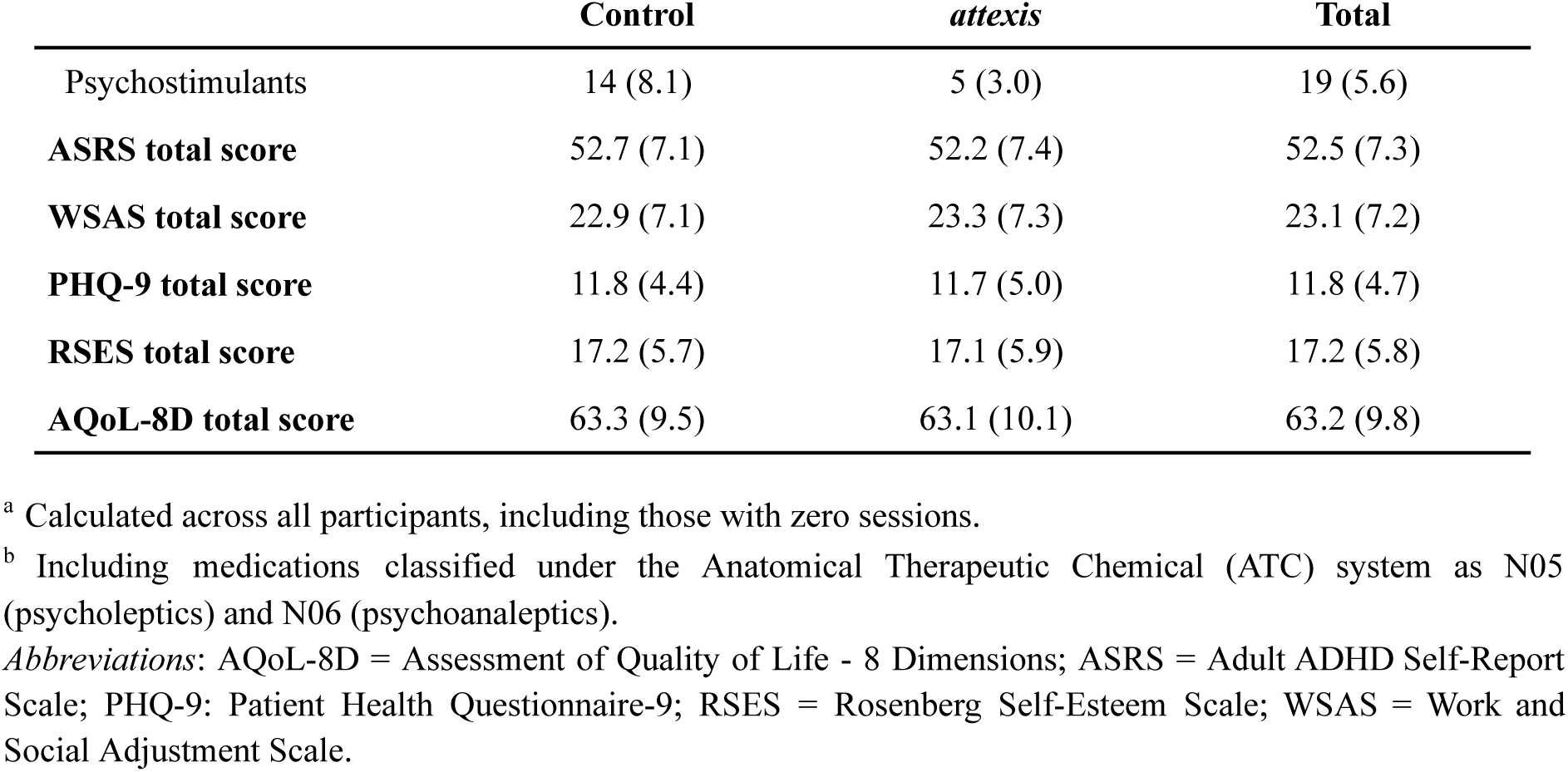
Subject demographics and clinical characteristics at baseline. Values represent mean (SD) unless stated otherwise.

### Missingness

Group differences in baseline characteristics between participants who completed the primary endpoint assessment at T1 (N = 316) and those who dropped out for any reason (N = 21) are reported in Supplementary Table 1. Compared to completers, dropouts were significantly younger, less likely to identify as White, more likely to be treated by an ADHD specialist, and more likely to be taking psychotropic medication. They also reported higher baseline levels of ADHD symptoms and functional impairment.

### Treatment effects

#### Primary endpoint: ADHD Symptom Severity

After 3 months, participants in the *attexis* + TAU group showed significantly lower ADHD symptom severity compared to the TAU-only group (see Table 3 and Figure 2). The baseline-adjusted mean difference on the ASRS total score was –5.0 points (95% CI = [–6.4, –3.6], *p* < .001; *d* = 0.85). Comparable effects were found in the J2R sensitivity analysis (–4.4 points, 95% CI [–5.7, –3.2], *p* < .001; *d* = 0.75) and the PP analysis (–5.0 points, 95% CI = [–6.4, –3.6], *p* < .001; *d* = 0.86). After 6 months, the ITT analysis confirmed a continued intervention effect (–4.5 points, 95% CI = [–6.2, –2.9], *p* < .001; *d* = 0.61). The J2R and PP analyses yielded comparable results (see Supplementary Tables 2 and 3), confirming the robustness of the treatment effect in relation to different assumptions concerning missing outcome data.

**Figure 2.**
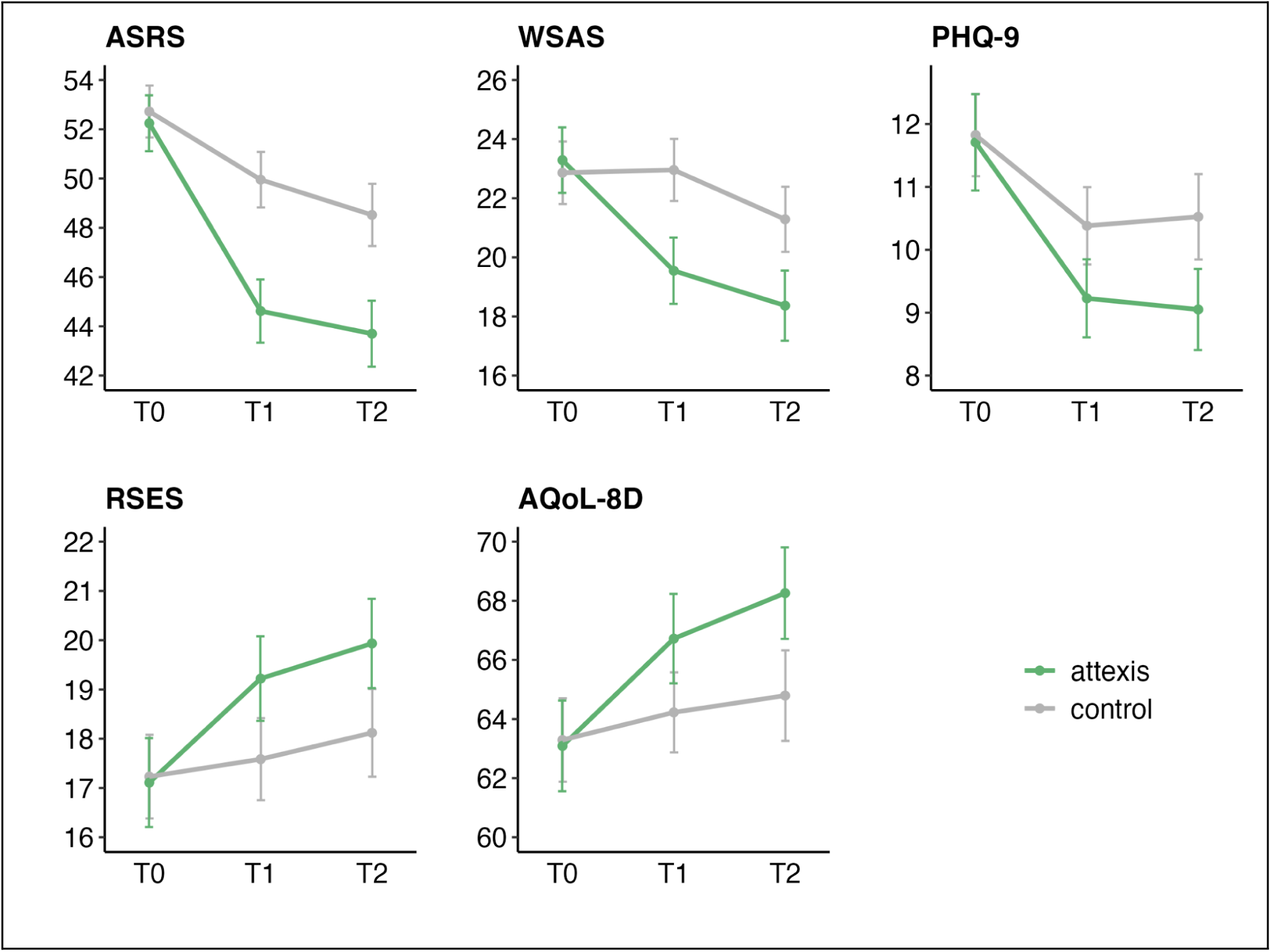
Symptom course on primary and secondary outcomes (means ± 95% confidence interval) using intent-to-treat analyses with multiple imputation for missing data. Plotted values represent total scores. *Abbreviations:* ASRS *=* Adult ADHD Self-Report Scale; AQoL-8D = Assessment of Quality of Life - 8 Dimensions; PHQ-9 = Patient Health Questionnaire-9; RSES = Rosenberg Self-Esteem Scale; WSAS = Work and Social Adjustment Scale

**Table 3.**
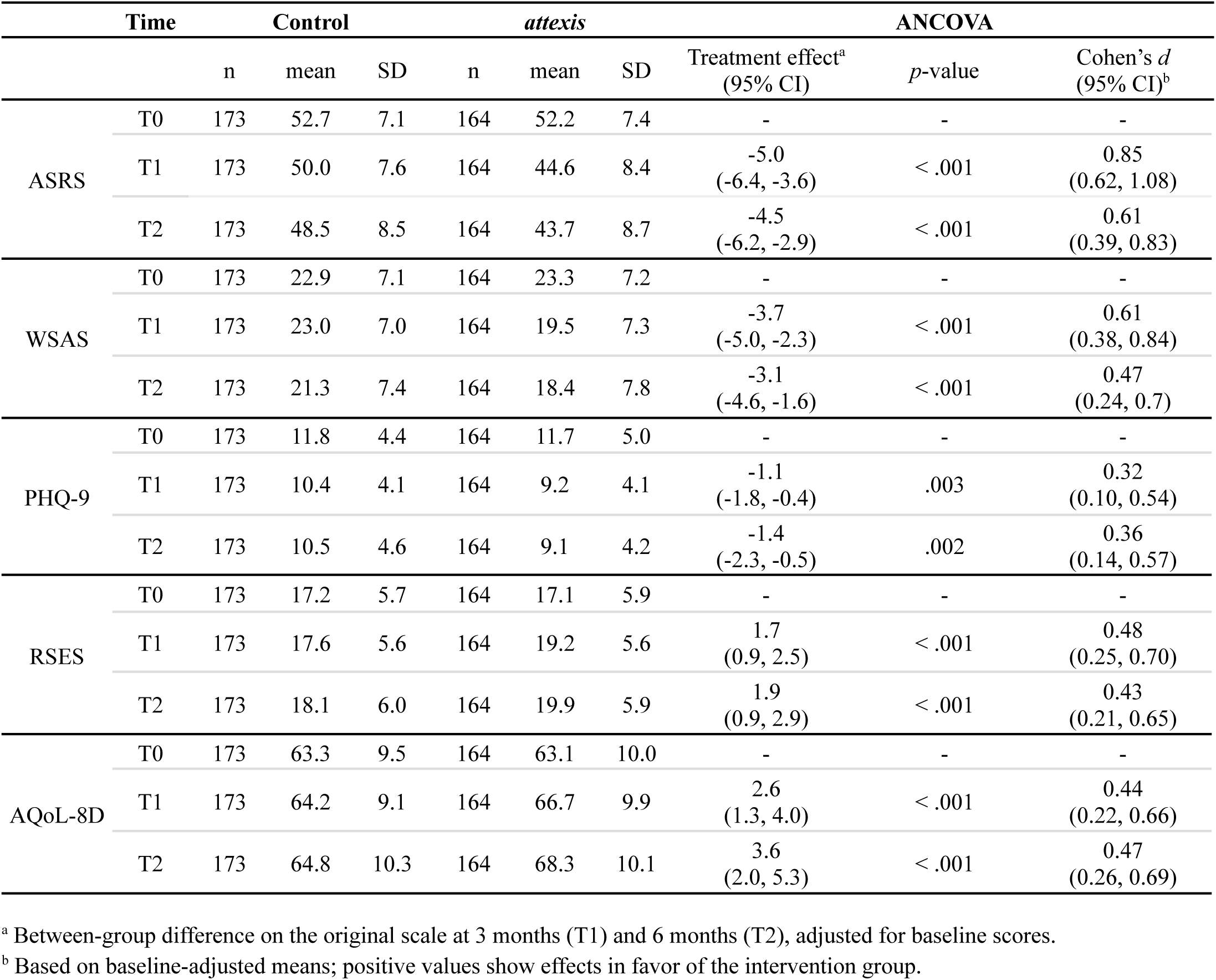
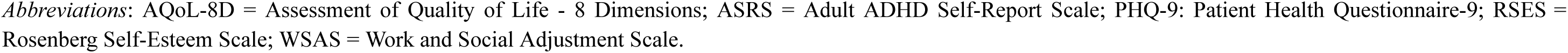
Results of primary and secondary endpoints for ITT analyses.

|  | Time | Control |  |  | *attaxis* |  |  | ANCOVA |  |  |
| --- | --- | --- | --- | --- | --- | --- | --- | --- | --- | --- |
|  |  | n | mean | SD | n | mean | SD | Treatment effect<sup>a</sup><br>(95% CI) | *p*-value | Cohen's *d*<br>(95% CI)<sup>b</sup> |
| ASRS | T0 | 173 | 52.7 | 7.1 | 164 | 52.2 | 7.4 | - | - | - |
|  | T1 | 173 | 50.0 | 7.6 | 164 | 44.6 | 8.4 | -5.0<br>(-6.4, -3.6) | < .001 | 0.85<br>(0.62, 1.08) |
|  | T2 | 173 | 48.5 | 8.5 | 164 | 43.7 | 8.7 | -4.5<br>(-6.2, -2.9) | < .001 | 0.61<br>(0.39, 0.83) |
| WSAS | T0 | 173 | 22.9 | 7.1 | 164 | 23.3 | 7.2 | - | - | - |
|  | T1 | 173 | 23.0 | 7.0 | 164 | 19.5 | 7.3 | -3.7<br>(-5.0, -2.3) | < .001 | 0.61<br>(0.38, 0.84) |
|  | T2 | 173 | 21.3 | 7.4 | 164 | 18.4 | 7.8 | -3.1<br>(-4.6, -1.6) | < .001 | 0.47<br>(0.24, 0.7) |
| PHQ-9 | T0 | 173 | 11.8 | 4.4 | 164 | 11.7 | 5.0 | - | - | - |
|  | T1 | 173 | 10.4 | 4.1 | 164 | 9.2 | 4.1 | -1.1<br>(-1.8, -0.4) | .003 | 0.32<br>(0.10, 0.54) |
|  | T2 | 173 | 10.5 | 4.6 | 164 | 9.1 | 4.2 | -1.4<br>(-2.3, -0.5) | .002 | 0.36<br>(0.14, 0.57) |
| RSES | T0 | 173 | 17.2 | 5.7 | 164 | 17.1 | 5.9 | - | - | - |
|  | T1 | 173 | 17.6 | 5.6 | 164 | 19.2 | 5.6 | 1.7<br>(0.9, 2.5) | < .001 | 0.48<br>(0.25, 0.70) |
|  | T2 | 173 | 18.1 | 6.0 | 164 | 19.9 | 5.9 | 1.9<br>(0.9, 2.9) | < .001 | 0.43<br>(0.21, 0.65) |
| AQoL-8D | T0 | 173 | 63.3 | 9.5 | 164 | 63.1 | 10.0 | - | - | - |
|  | T1 | 173 | 64.2 | 9.1 | 164 | 66.7 | 9.9 | 2.6<br>(1.3, 4.0) | < .001 | 0.44<br>(0.22, 0.66) |
|  | T2 | 173 | 64.8 | 10.3 | 164 | 68.3 | 10.1 | 3.6<br>(2.0, 5.3) | < .001 | 0.47<br>(0.26, 0.69) |

<sup>a</sup> Between-group difference on the original scale at 3 months (T1) and 6 months (T2), adjusted for baseline scores.
<sup>b</sup> Based on baseline-adjusted means; positive values show effects in favor of the intervention group.
*Abbreviations:* AQoL-8D = Assessment of Quality of Life - 8 Dimensions; ASRS = Adult ADHD Self-Report Scale; PHQ-9: Patient Health Questionnaire-9; RSES = Rosenberg Self-Esteem Scale; WSAS = Work and Social Adjustment Scale.

After 3 months, 11.6% of participants in the intervention group (19/164) met the predefined responder criterion (≥ 30% reduction in the ASRS total score), compared to 1.2% in the control group (2/173). The difference between groups was statistically significant (χ² = 15.67, *p* < .001), corresponding to an OR of 11.2 (95% CI [2.6, 48.9]). The safety analysis further indicated that symptom worsening (i.e., any increase in ASRS total score from baseline to T1) was observed in 10.7% of participants in the intervention group (16/149) and 24% in the control group (40/167). The difference was statistically significant (χ² = 9.43, *p* = .002; OR = 2.6, 95% CI [1.4, 5.0]), indicating that the use of *attexis* was associated with a lower likelihood of symptom worsening compared to TAU alone.

#### Secondary endpoints

After 3 months, ITT analyses showed significant improvements in the intervention group across all secondary outcomes (see Table 3 and Figure 2), including work and social functioning, depression, self-esteem, and quality of life. Effect sizes ranged from *d* = 0.32 to 0.61. J2R and PP analyses yielded consistent results (see Supplementary Tables 2 and 3).

Responder analyses confirmed the clinical relevance of these effects: significantly more participants in the intervention group showed meaningful improvements in work and social functioning (based on an MCID of 8 points (Zahra et al., 2014); 26.2% vs. 11.6%; χ² = 11.9, *p* < .001; OR = 2.7, 95% CI [1.5, 4.9]), depression levels (based on an MCID of 5 points (Löwe, Unützer, Callahan, Perkins, & Kroenke, 2004); 31.1% vs. 19.7%; χ² = 5.85, *p* = .016; OR = 1.8, 95% CI [1.1, 3.0]), self-esteem (based on the RCI; 20.1% vs. 9.8%; χ² = 7.06, *p* = .008; OR = 2.3, 95% CI [1.2, 4.3]), and quality of life (based on the RCI; 21.3% vs. 9.8%; χ² = 8.55, *p* = .003; OR = 2.5, 95% CI [1.3, 4.6]).

At 6 months, intervention effects continued to be observable across all secondary outcomes (see Table 3 and Figure 2). Effect sizes ranged from *d* = 0.36 to 0.47. These results were consistent in both PP and J2R analyses (see Supplementary Tables 2 and 3).

#### Subgroup analyses

Subgroup analyses showed consistent effectiveness of *attexis* across sex, psychotherapy status at baseline, psychotropic medication use, and treatment changes between T0 and T1 (see Supplementary Tables 4–7).

#### User satisfaction

Mean ratings for the likelihood to recommend the program to a friend or colleague were 6.1 (SD = 2.9) at T1 and 5.9 (SD = 3.3) at T2, indicating a generally positive evaluation of *attexis*.

Subjective ADHD symptom improvement (assessed via the PGIC) was significantly greater in the intervention group at both T1 and T2 (T1: *d* = 0.56; T2: *d* = 0.55; both *ps* < .001). Similar effects were observed for perceived improvements in daily functioning (T1: *d* = 0.66; T2: *d* = 0.63; both *ps* < .001). In binary ratings, 36.5% of participants in the intervention group reported meaningful perceived functional improvement at T1 vs. 9.6% in the control group (χ² = 32.9, *p* < .001); at T2, this was 40.1% vs. 17.6% (χ² = 18.8, *p* < .001).

#### Adverse events

After 3 months, unplanned or emergency medical treatments were reported by 7.8% of participants in the control group (13/167) and 5.4% in the intervention group (8/147), with no significant group difference (χ² = 0.69, *p* = .407). A similar pattern was observed after 6 months (control: 6.3%, 10/159; intervention: 4.2%, 6/142; χ² = 0.63, *p* = .426). No adverse events were linked to the use of *attexis*, and no adverse device effects were reported.

## Discussion

This pragmatic RCT evaluated the effectiveness of *attexis*, a fully self-guided digital intervention based on cognitive-behavioral and mindfulness principles for adults with ADHD. Results show that *attexis,* when added to TAU, significantly reduced ADHD symptom severity and improved a range of patient-relevant functional and psychosocial secondary outcomes compared to TAU-only. Effects were robust under conservative assumptions regarding missing outcome data and sustained at the 6-month follow-up, highlighting the long-term potential of the intervention. Responder analyses confirmed the clinical relevance of the intervention effects across all outcomes. Participants reported good satisfaction with *attexis*, along with perceived improvements in ADHD symptoms and their impact on daily life. Importantly, no adverse events were attributed to the use of *attexis*.

In terms of the primary outcome, ADHD symptom severity after 3 months, *attexis* demonstrated a large effect size (*d* = 0.85). This finding aligns with previous research on digital interventions for adult ADHD, which reported effect sizes ranging from *d* = 0.42 to 1.21 for this outcome (Kenter et al., 2023; Moëll et al., 2015; Nasri et al., 2023; Pettersson et al., 2017; Selaskowski et al., 2022). However, all previous studies featured small sample sizes, a common limitation in research on non-pharmacological treatments for adult ADHD (Nimmo-Smith et al., 2020). In contrast, the present study was adequately powered and, to our knowledge, represents the largest RCT to date evaluating a digital intervention for adults with ADHD. Therefore, our study strengthens the evidence base for digital interventions in this population. Notably, the magnitude of the intervention effect falls within the range reported in meta-analyses of face-to-face CBT for adult ADHD (*d* = 0.71-0.98) (Liu et al., 2023; Lopez et al., 2018; Young et al., 2020). Thus, our results mirror previous meta-analytic findings suggesting that therapeutic content may matter more than delivery format in the therapy of adult ADHD (Knouse et al., 2017).

Regarding secondary outcomes, use of *attexis* was linked to consistent benefits in a wide range of functional and psychosocial domains, specifically, occupational and social functioning, depressive symptoms, self-esteem, and quality of life. These outcomes are highly relevant from a patient perspective, as they reflect the real-world burden of ADHD and are closely linked to long-term functioning and well-being (Coghill, Banaschewski, Soutullo, Cottingham, & Zuddas, 2017; Cook, Knight, Hume, & Qureshi, 2014; D’Amelio, Retz, Philipsen, & Rösler, 2009). Despite their clinical relevance and inclusion in guideline recommendations, they remain insufficiently addressed in both routine care and research (AWMF, 2017; Kooij, Bijlenga, et al., 2019; López-Pinar, Martínez-Sanchís, Carbonell-Vayá, Sánchez-Meca, & Fenollar-Cortés, 2020). This may, in part, be due to the fact that pharmacotherapy, currently the predominant treatment approach for adult ADHD, has shown only limited and inconsistent effects on these broader psychosocial outcome domains (Bellato et al., 2024; Castells, Blanco-Silvente, & Cunill, 2018; Coghill et al., 2017; Lenzi, Cortese, Harris, & Masi, 2018).

Over the 6-month follow-up, we observed relevant dynamics in the evaluated outcomes: While the intervention effects of *attexis* on core ADHD symptoms and functional impairment were maintained at 6 months, improvements in depressive symptoms and quality of life increased further over time. This pattern suggests that as core symptoms decrease, individuals begin to engage more with their environment and implement adaptive coping strategies in daily life. According to CBT models of adult ADHD, such behavioral engagement and cognitive restructuring can, over time, reduce comorbid internalizing symptoms and the broader real-world burden associated with ADHD (López-Pinar et al., 2020; Safren, Sprich, Chulvick, & Otto, 2004).

Overall, our findings address a relevant gap in the current treatment for adults with ADHD. Psychosocial interventions, particularly CBT, are recommended in treatment of adult ADHD (AWMF, 2017; Kooij, Bijlenga, et al., 2019), but access remains limited, with long waiting times and a lack of trained professionals representing well-documented barriers in Germany (Kruse et al., 2024; Libutzki et al., 2019; Schneider et al., 2023). This is reflected in our sample, where only 3.6% of participants received care by an ADHD specialist at baseline. Importantly, many adults with ADHD report that pharmacological treatment alone is inadequate and express a desire for psychoeducation and psychosocial support (AWMF, 2017; Matheson et al., 2013). Reflecting this, meta-analytic evidence indicates that combining pharmacotherapy with CBT may lead to improved treatment outcomes in adult ADHD compared to pharmacotherapy alone (Lopez et al., 2018). Easily accessible digital interventions such as *attexis* may represent a valuable addition to the treatment of adult ADHD across different clinical contexts, including, as evidenced by subgroup analyses, both as a stand-alone tool and combination with pharmacotherapy.

### Limitations

Several limitations need to be considered when interpreting our results. First, while ADHD diagnoses were confirmed before inclusion by trained clinicians using structured interviews, all outcome measures were based on self-report, which may be subject to recall or social desirability bias. In line with patient-centered care, however, self-report captures patients’ subjective experience and the use of validated instruments and objective responder criteria supports the reliability of the findings. Second, participants were not blinded to group allocation, which may have exaggerated intervention effects via expectancy effects. That said, blinding is generally difficult in psychotherapy research, as the development of credible but ineffective sham interventions poses significant conceptual challenges (Kirsch, Wampold, & Kelley, 2016). Third, satisfaction ratings suggest that experiences with the program varied. Future qualitative research should examine more in-depth which components were perceived as helpful or lacking by patients. Fourth, attrition was overall low compared to the literature (Linardon & Fuller-Tyszkiewicz, 2020), but slightly higher in the intervention group. While this pattern is common (Assmann et al., 2025; Betz, Jacob, Knitza, Koehm, & Behrens, 2024; Specht et al., 2024) and was addressed analytically, underlying reasons for dropout warrant further investigation. Finally, generalizability may be limited to digitally savvy individuals willing to engage with a digital solution for treatment of ADHD symptoms. However, this population likely mirrors those who are most likely to use and benefit from prescribed digital interventions in real-world care settings (Betz et al., 2024).

### Conclusion

In conclusion, this study adds important evidence to the growing field of digital interventions for adult ADHD. *attexis*, a fully self-guided digital intervention based on CBT and mindfulness principles, was associated with significant and clinically relevant reductions in ADHD symptom severity and improvements in a broad range of patient-relevant secondary outcomes, including functional impairment, depressive symptoms, self-esteem and quality of life. The intervention showed consistent effects across diverse subgroups and was well accepted by users. While further research is needed to replicate and extend these findings, particularly in routine care settings, digital interventions like *attexis* may offer a promising treatment option for adults with ADHD, especially in light of limited access to evidence-based psychosocial interventions.

## Data Availability

The data produced in the present study are not publicly available due to proprietary reasons.

## Acknowledgements

The authors would like to thank all patients who took part in the READ-ADHD RCT.

## Required statements

### Ethical standards

The authors assert that all procedures contributing to this work comply with the ethical standards of the relevant national and institutional committees on human experimentation and with the Helsinki Declaration of 1975, as revised in 2008.

### Author note

Part of these findings were presented by LTB at the 2024 Annual Congress of the German Association for Psychiatry, Psychotherapy and Psychosomatics (DGPPN) in Berlin, Germany.

### Contributorship

Conception and design of the study: RD, LTB, JPK, GAJ, PRJ

Data collection: LTB, SMJ, GAJ

Data analysis and interpretation: LTB, RD, PR, GAJ

Drafting of the manuscript: LTB, RD, GAJ, PRJ

Critical revision of the manuscript for important intellectual content: RD, LTB, SMJ, WR, AP, JPK, EF, GAJ, PRJ

Final approval of the version to be published: RD, LTB, SMJ, WR, AP, JPK, EF, GAJ, PRJ

## References

Andersson, G. (2016). Internet-Delivered Psychological Treatments. Annual Review of Clinical Psychology, 12(1), 157–179. 10.1146/annurev-clinpsy-021815-093006

Assmann, N., Jacob, G. A., Schaich, A., Berger, T., Zindler, T., Betz, L., … Klein, J. P. (2025). A digital therapeutic for people with borderline personality disorder: A pragmatic, single blind, randomised controlled trial. Lancet Psychiatry, 12(5), 366–376.

AWMF. (2017). *Langfassung der interdisziplinären evidenz- und konsensbasierten (S3) Leitlinie “Aufmerksamkeitsdefizit-/Hyperaktivitätsstörung (ADHS) im Kindes-, Jugend- und Erwachsenenalter”* (Association of the Scientific Medical Societies in Germany, Ed.). Retrieved from https://register.awmf.org/assets/guidelines/028-045l_S3_ADHS_2018-06-abgelaufen.pdf

Bartlett, J. W. (2023). Reference-Based Multiple Imputation–What is the Right Variance and How to Estimate It. Statistics in Biopharmaceutical Research, 15(1), 178–186. 10.1080/19466315.2021.1983455

Bellato, A., Perrott, N. J., Marzulli, L., Parlatini, V., Coghill, D., & Cortese, S. (2024). Systematic Review and Meta-Analysis: Effects of Pharmacological Treatment for Attention-Deficit/Hyperactivity Disorder on Quality of Life. Journal of the American Academy of Child & Adolescent Psychiatry. 10.1016/j.jaac.2024.05.023

Betz, L. T., Jacob, G. A., Knitza, J., Koehm, M., & Behrens, F. (2024). Efficacy of a cognitive-behavioral digital therapeutic on psychosocial outcomes in rheumatoid arthritis: Randomized controlled trial. Npj Mental Health Research, 3(1), 1–11. 10.1038/s44184-024-00085-8

Buitelaar, J. K., Montgomery, S. A., & van Zwieten-Boot, B. J. (2003). Attention deficit hyperactivity disorder: Guidelines for investigating efficacy of pharmacological intervention. European Neuropsychopharmacology, 13(4), 297–304. 10.1016/S0924-977X(03)00047-6

Castells, X., Blanco-Silvente, L., & Cunill, R. (2018). Amphetamines for attention deficit hyperactivity disorder (ADHD) in adults. The Cochrane Database of Systematic Reviews, 8(8), CD007813. 10.1002/14651858.CD007813.pub3

Coghill, D. R., Banaschewski, T., Soutullo, C., Cottingham, M. G., & Zuddas, A. (2017). Systematic review of quality of life and functional outcomes in randomized placebo-controlled studies of medications for attention-deficit/hyperactivity disorder. European Child & Adolescent Psychiatry, 26(11), 1283–1307. 10.1007/s00787-017-0986-y

Cook, J., Knight, E., Hume, I., & Qureshi, A. (2014). The self-esteem of adults diagnosed with attention-deficit/hyperactivity disorder (ADHD): A systematic review of the literature. Attention Deficit and Hyperactivity Disorders, 6(4), 249–268. 10.1007/s12402-014-0133-2

D’Amelio, R., Retz, W., Philipsen, A., & Rösler, M. (Eds.). (2009). *Psychoedukation und Coaching: ADHS im Erwachsenenalter; Manual zur Leitung von Patienten- und Angehörigengruppen* (1st ed.). München, Jena: Elsevier, Urban & Fischer.

de Zwaan, M., Gruß, B., Müller, A., Graap, H., Martin, A., Glaesmer, H., … Philipsen, A. (2012). The estimated prevalence and correlates of adult ADHD in a German community sample. European Archives of Psychiatry and Clinical Neuroscience, 262(1), 79–86. 10.1007/s00406-011-0211-9

Faraone, S. V., Bellgrove, M. A., Brikell, I., Cortese, S., Hartman, C. A., Hollis, C., … Buitelaar, J. K. (2024). Attention-deficit/hyperactivity disorder. Nature Reviews Disease Primers, 10(1), 1–21. 10.1038/s41572-024-00495-0

Fayyad, J., Sampson, N. A., Hwang, I., Adamowski, T., Aguilar-Gaxiola, S., Al-Hamzawi, A., … WHO World Mental Health Survey Collaborators. (2017). The descriptive epidemiology of DSM-IV Adult ADHD in the World Health Organization World Mental Health Surveys. Attention Deficit and Hyperactivity Disorders, 9(1), 47–65. 10.1007/s12402-016-0208-3

Fuermaier, A. B. M., Tucha, L., Butzbach, M., Weisbrod, M., Aschenbrenner, S., & Tucha, O. (2021). ADHD at the workplace: ADHD symptoms, diagnostic status, and work-related functioning. Journal of Neural Transmission, 128(7), 1021–1031. 10.1007/s00702-021-02309-z

Guy, W. (1976). Clinical global impression. Assessment Manual for Psychopharmacology, 217–222. 10.1037/t48216-000

Heissel, A., Bollmann, J., Kangas, M., Abdulla, K., Rapp, M., & Sanchez, A. (2021). Validation of the German version of the work and social adjustment scale in a sample of depressed patients. BMC Health Services Research, 21(1), 593. 10.1186/s12913-021-06622-x

Hennig, T., Koglin, U., Schmidt, S., Petermann, F., & Brähler, E. (2017). Attention-deficit/hyperactivity disorder symptoms and life satisfaction in a representative adolescent and adult sample. The Journal of Nervous and Mental Disease, 205(9), 720–724. 10.1097/NMD.0000000000000700

Hoxhaj, E., Sadohara, C., Borel, P., D’Amelio, R., Sobanski, E., Müller, H., … Philipsen, A. (2018). Mindfulness vs psychoeducation in adult ADHD: A randomized controlled trial. European Archives of Psychiatry and Clinical Neuroscience, 268(4), 321–335. 10.1007/s00406-018-0868-4

Karyotaki, E., Efthimiou, O., Miguel, C., genannt Bermpohl, F. M., Furukawa, T. A., Cuijpers, P., … Gemmil, A. W. (2021). Internet-based cognitive behavioral therapy for depression: A systematic review and individual patient data network meta-analysis. JAMA Psychiatry, 78(4), 361–371. 10.1001/jamapsychiatry.2020.4364

Kenter, R. M. F., Gjestad, R., Lundervold, A. J., & Nordgreen, T. (2023). A self-guided internet-delivered intervention for adults with ADHD: Results from a randomized controlled trial. Internet Interventions, 32, 100614. 10.1016/j.invent.2023.100614

Kessler, R. C., Adler, L., Ames, M., Demler, O., Faraone, S., Hiripi, E., … Walters, E. E. (2005). The World Health Organization adult ADHD self-report scale (ASRS): A short screening scale for use in the general population. Psychological Medicine, 35(2), 245–256. 10.1017/S0033291704002892

Kirsch, I., Wampold, B. E., & Kelley, J. M. (2016). Controlling for the placebo effect in psychotherapy: Noble quest or tilting at windmills? *Psychology of Consciousness: Theory*, Research, and Practice, 3(2), 121–131. 10.1037/cns0000065

Kliem, S., Sachser, C., Lohmann, A., Baier, D., Braehler, E., Gündel, H., & Fegert, J. M. (2024). Psychometric evaluation and community norms of the PHQ-9, based on a representative German sample. Frontiers in Psychiatry, 15, 1483782. 10.3389/fpsyt.2024.1483782

Knouse, L. E., Teller, J., & Brooks, M. A. (2017). Meta-analysis of cognitive–behavioral treatments for adult ADHD. Journal of Consulting and Clinical Psychology, 85(7), 737. 10.1037/ccp0000216

Koerts, J., Bangma, D. F., Fuermaier, A. B. M., Mette, C., Tucha, L., & Tucha, O. (2021). Financial judgment determination in adults with ADHD. Journal of Neural Transmission, 128(7), 969–979. 10.1007/s00702-021-02323-1

Kooij, J. J. S., Bijlenga, D., Salerno, L., Jaeschke, R., Bitter, I., Balazs, J., … others. (2019). Updated European Consensus Statement on diagnosis and treatment of adult ADHD. European Psychiatry, 56(1), 14–34. 10.1016/j.eurpsy.2018.11.001

Kooij, J. J. S., Francken, M. H., Bron, A., & Wynchank, D. (2019). Diagnostisches Interview für ADHS bei Erwachsenen – DIVA-5. Retrieved from https://www.divacenter.eu

Kruse, J., Kampling, H., Bouami, S. F., Grobe, T. G., Hartmann, M., Jedamzik, J., … Friederich, H.-C. (2024). Outpatient Psychotherapy in Germany. Deutsches Ärzteblatt International, 121(10), 315–322. 10.3238/arztebl.m2024.0039

Lenzi, F., Cortese, S., Harris, J., & Masi, G. (2018). Pharmacotherapy of emotional dysregulation in adults with ADHD: A systematic review and meta-analysis. Neuroscience & Biobehavioral Reviews, 84, 359–367. 10.1016/j.neubiorev.2017.08.010

Libutzki, B., Ludwig, S., May, M., Jacobsen, R. H., Reif, A., & Hartman, C. A. (2019). Direct medical costs of ADHD and its comorbid conditions on basis of a claims data analysis. European Psychiatry, 58, 38–44. 10.1016/j.eurpsy.2019.01.019

Linardon, J., & Fuller-Tyszkiewicz, M. (2020). Attrition and adherence in smartphone-delivered interventions for mental health problems: A systematic and meta-analytic review. Journal of Consulting and Clinical Psychology, 88(1), 1–13. 10.1037/ccp0000459

Liu, C.-I., Hua, M.-H., Lu, M.-L., & Goh, K. K. (2023). Effectiveness of cognitive behavioural-based interventions for adults with attention-deficit/hyperactivity disorder extends beyond core symptoms: A meta-analysis of randomized controlled trials. Psychology and Psychotherapy: Theory, Research and Practice, 96(3), 543–559. 10.1111/papt.12455

Lopez, P. L., Torrente, F. M., Ciapponi, A., Lischinsky, A. G., Cetkovich-Bakmas, M., Rojas, J. I., … Manes, F. F. (2018). Cognitive-behavioural interventions for attention deficit hyperactivity disorder (ADHD) in adults. Cochrane Database of Systematic Reviews, (3). 10.1002/14651858.CD010840.pub2

López-Pinar, C., Martínez-Sanchís, S., Carbonell-Vayá, E., Sánchez-Meca, J., & Fenollar-Cortés, J. (2020). Efficacy of nonpharmacological treatments on comorbid internalizing symptoms of adults with attention-deficit/hyperactivity disorder: A meta-analytic review. Journal of Attention Disorders, 24(3), 456–478. 10.1177/1087054719855685

Löwe, B., Unützer, J., Callahan, C. M., Perkins, A. J., & Kroenke, K. (2004). Monitoring depression treatment outcomes with the Patient Health Questionnaire-9. Medical Care, 42(12), 1194–1201. 10.1097/00005650-200412000-00006

Lundqvist, J., Lindberg, M. S., Brattmyr, M., Havnen, A., Hjemdal, O., & Solem, S. (2024). The Work and Social Adjustment Scale (WSAS): An investigation of reliability, validity, and associations with clinical characteristics in psychiatric outpatients. Plos One, 19(10), e0311420. 10.1371/journal.pone.0311420

Margraf, J., Cwik, J. C., Pflug, V., & Schneider, S. (2017). Strukturierte klinische Interviews zur Erfassung psychischer Störungen über die Lebensspanne. Zeitschrift Für Klinische Psychologie Und Psychotherapie, 46(3), 176–186. 10.1026/1616-3443/a000430

Martin, A., Rief, W., Klaiberg, A., & Braehler, E. (2006). Validity of the Brief Patient Health Questionnaire Mood Scale (PHQ-9) in the general population. General Hospital Psychiatry, 28(1), 71–77. 10.1016/j.genhosppsych.2005.07.003

Masuch, T. V., Bea, M., Alm, B., Deibler, P., & Sobanski, E. (2019). Internalized stigma, anticipated discrimination and perceived public stigma in adults with ADHD. ADHD Attention Deficit and Hyperactivity Disorders, 11, 211–220. 10.1007/s12402-018-0274-9

Matheson, L., Asherson, P., Wong, I. C. K., Hodgkins, P., Setyawan, J., Sasane, R., & Clifford, S. (2013). Adult ADHD patient experiences of impairment, service provision and clinical management in England: A qualitative study. BMC Health Services Research, 13, 184. 10.1186/1472-6963-13-184

Moëll, B., Kollberg, L., Nasri, B., Lindefors, N., & Kaldo, V. (2015). Living SMART — A randomized controlled trial of a guided online course teaching adults with ADHD or sub-clinical ADHD to use smartphones to structure their everyday life. Internet Interventions, 2(1), 24–31. 10.1016/j.invent.2014.11.004

Mörstedt, B., Corbisiero, S., & Stieglitz, R.-D. (2016). Normierung der Adult ADHD Self-Report-Scale-V1.1 und der ADHS Selbstbeurteilungsskala an einer repräsentativen deutschsprachigen Stichprobe. Diagnostica, 62(4), 199–211. 10.1026/0012-1924/a000154

Mundt, J. C., Marks, I. M., Shear, M. K., & Greist, J. H. (2002). The Work and Social Adjustment Scale: A simple measure of impairment in functioning. The British Journal of Psychiatry: The Journal of Mental Science, 180, 461–464. 10.1192/bjp.180.5.461

Nasri, B., Cassel, M., Enhärje, J., Larsson, M., Hirvikoski, T., Ginsberg, Y., … Kaldo, V. (2023). Internet delivered cognitive behavioral therapy for adults with ADHD-A randomized controlled trial. Internet Interventions, 33, 100636. 10.1016/j.invent.2023.100636

NICE. (2018). Attention deficit hyperactivity disorder: Diagnosis and management. London. Retrieved from http://www.ncbi.nlm.nih.gov/books/NBK493361/

Nimmo-Smith, V., Merwood, A., Hank, D., Brandling, J., Greenwood, R., Skinner, L., … Rai, D. (2020). Non-pharmacological interventions for adult ADHD: A systematic review. Psychological Medicine, 50(4), 529–541. 10.1017/S0033291720000069

Pauley, D., Cuijpers, P., Papola, D., Miguel, C., & Karyotaki, E. (2023). Two decades of digital interventions for anxiety disorders: A systematic review and meta-analysis of treatment effectiveness. Psychological Medicine, 53(2), 567–579. 10.1017/S0033291721001999

Pettersson, R., Söderström, S., Edlund-Söderström, K., & Nilsson, K. W. (2017). Internet-Based Cognitive Behavioral Therapy for Adults With ADHD in Outpatient Psychiatric Care. Journal of Attention Disorders, 21(6), 508–521. 10.1177/1087054714539998

Philipsen, A., & Döpfner, M. (2020). ADHS im Übergang in das Erwachsenenalter: Prävalenz, Symptomatik, Risiken und Versorgung. Bundesgesundheitsblatt - Gesundheitsforschung - Gesundheitsschutz, 63(7), 910–915. 10.1007/s00103-020-03175-y

Philipsen, A., Jans, T., Graf, E., Matthies, S., Borel, P., Colla, M., … for the Comparison of Methylphenidate and Psychotherapy in Adult ADHD Study (COMPAS) Consortium. (2015). Effects of Group Psychotherapy, Individual Counseling, Methylphenidate, and Placebo in the Treatment of Adult Attention-Deficit/Hyperactivity Disorder: A Randomized Clinical Trial. JAMA Psychiatry, 72(12), 1199–1210. 10.1001/jamapsychiatry.2015.2146

R Core Team. (2024). R: A language and environment for statistical computing. Vienna, Austria. Retrieved from https://cran.r-project.org/

Richardson, J., Iezzi, A., Khan, M. A., & Maxwell, A. (2014). Validity and reliability of the Assessment of Quality of Life (AQoL)-8D multi-attribute utility instrument. The Patient, 7(1), 85–96. 10.1007/s40271-013-0036-x

Richardson, J., Khan, M. A., Iezzi, A., & Maxwell, A. (2012). Cross-national comparison of twelve quality of life instruments. MIC Paper, 2. Retrieved from https://www.aqol.com.au/papers/researchpaper85.pdf

Rosenberg, M. (1965). Rosenberg self-esteem scale (RSE). Princeton, New Jersey: Princeton University Press.

Roth, M., Decker, O., Herzberg, P. Y., & Brähler, E. (2008). Dimensionality and norms of the Rosenberg Self-Esteem Scale in a German general population sample. European Journal of Psychological Assessment, 24(3), 190–197. 10.1027/1015-5759.24.3.190

Safren, S. A., Sprich, S., Chulvick, S., & Otto, M. W. (2004). Psychosocial treatments for adults with attention-deficit/hyperactivity disorder. Psychiatric Clinics, 27(2), 349–360. 10.1016/S0193-953X(03)00089-3

Schlander, M., Schwarz, O., Trott, G.-E., Viapiano, M., & Bonauer, N. (2007). Who cares for patients with attention-deficit/hyperactivity disorder (ADHD)? European Child & Adolescent Psychiatry, 16(7), 430–438. 10.1007/s00787-007-0616-1

Schneider, B. C., Schöttle, D., Hottenrott, B., Gallinat, J., & Moritz, S. (2023). Assessment of Adult ADHD in Clinical Practice: Four Letters—40 Opinions. Journal of Attention Disorders, 27(9), 1051–1061. 10.1177/1087054719879498

Selaskowski, B., Steffens, M., Schulze, M., Lingen, M., Aslan, B., Rosen, H., … Boll, S. (2022). Smartphone-assisted psychoeducation in adult attention-deficit/hyperactivity disorder: A randomized controlled trial. Psychiatry Research, 317, 114802.

Simon, V., Czobor, P., Bálint, S., Mészáros, A., & Bitter, I. (2009). Prevalence and correlates of adult attention-deficit hyperactivity disorder: Meta-analysis. The British Journal of Psychiatry, 194(3), 204–211.

Song, P., Zha, M., Yang, Q., Zhang, Y., Li, X., & Rudan, I. (2021). The prevalence of adult attention-deficit hyperactivity disorder: A global systematic review and meta-analysis. Journal of Global Health, 11, 04009. 10.7189/jogh.11.04009

Specht, A., Betz, L. T., Riepenhausen, A., Jauch-Chara, K., Jacob, G. A., Riemann, D., & Göder, R. (2024). Effectiveness and safety of an interactive internet-based intervention to improve insomnia: Results from a randomised controlled trial. Journal of Sleep Research, e14409. 10.1111/jsr.14409

Thorell, L. B., Holst, Y., & Sjöwall, D. (2019). Quality of life in older adults with ADHD: Links to ADHD symptom levels and executive functioning deficits. Nordic Journal of Psychiatry, 73(7), 409–416. 10.1080/08039488.2019.1646804

von Collani, G., & Herzberg, P. Y. (2003). Eine revidierte Fassung der deutschsprachigen Skala zum Selbstwertgefühl von Rosenberg. [A revised version of the German adaptation of Rosenberg’s Self-Esteem Scale.]. Zeitschrift Für Differentielle Und Diagnostische Psychologie, 24, 3–7. 10.1024/0170-1789.24.1.3

von Hippel, P. T., & Bartlett, J. W. (2021). Maximum likelihood multiple imputation: Faster imputations and consistent standard errors without posterior draws. Statistical Science, 36(3), 400–420. 10.1214/20-STS793

Young, Z., Moghaddam, N., & Tickle, A. (2020). The Efficacy of Cognitive Behavioral Therapy for Adults With ADHD: A Systematic Review and Meta-Analysis of Randomized Controlled Trials. Journal of Attention Disorders, 24(6), 875–888. 10.1177/1087054716664413

Zahra, D., Qureshi, A., Henley, W., Taylor, R., Quinn, C., Pooler, J., … Byng, R. (2014). The work and social adjustment scale: Reliability, sensitivity and value. International Journal of Psychiatry in Clinical Practice, 18(2), 131–138. 10.3109/13651501.2014.894072

